# In vitro, classical complement activation differs by disease severity and between SARS-CoV-2 antigens

**DOI:** 10.1101/2021.11.22.21266681

**Authors:** Rachel E Lamerton, Edith Marcial-Juarez, Sian E Faustini, Marisol Perez-Toledo, Margaret Goodall, Siân E Jossi, Maddy L Newby, Iain Chapple, Thomas Dietrich, Tonny Veenith, Adrian M Shields, Lorraine Harper, Ian R Henderson, Julie Rayes, David C Wraith, Steve P Watson, Max Crispin, Mark T Drayson, Alex G Richter, Adam F Cunningham

## Abstract

Antibodies specific for the spike glycoprotein (S) and nucleocapsid (N) SARS-CoV-2 proteins are typically present during severe COVID-19, and induced to S after vaccination. The binding of viral antigens by antibody can initiate the classical complement pathway. Since complement could play pathological or protective roles at distinct times during SARS-CoV-2 infection we determined levels of antibody-dependent complement activation along the complement cascade. Here, we used an ELISA assay to assess complement protein binding (C1q) and the deposition of C4b, C3b, and C5b to S and N antigens in the presence of anti-SARS-CoV-2 antibodies from different test groups: non-infected, single and double vaccinees, non-hospitalised convalescent (NHC) COVID-19 patients and convalescent hospitalised (ITU-CONV) COVID-19 patients. C1q binding correlates strongly with antibody responses, especially IgG1 levels. However, detection of downstream complement components, C4b, C3b and C5b shows some variability associated with the antigen and subjects studied. In the ITU-CONV, detection of C3b-C5b to S was observed consistently, but this was not the case in the NHC group. This is in contrast to responses to N, where median levels of complement deposition did not differ between the NHC and ITU-CONV groups. Moreover, for S but not N, downstream complement components were only detected in sera with higher IgG1 levels. Therefore, the classical pathway is activated by antibodies to multiple SARS-CoV-2 antigens, but the downstream effects of this activation may differ depending on the specific antigen targeted and the disease status of the subject.

- **Spike- and nucleocapsid-specific antibodies activate complement *in vitro***
- **C1q binding correlates with IgG1 antibody levels**
- **Generation of C4b, C3b and C5b relates to the antigen targeted and the patient group tested**

## Introduction

Infection with SARS-CoV-2, the causative agent of COVID-19, results in a spectrum of clinical presentations ranging from asymptomatic infections to severe disease and death. Although some factors that can predict risk of severe disease are known, such as obesity or age, it is clear that other host factors, including immune status, also contribute [1-3]. Thus, it is likely that COVID-19 represents a collection of syndromes, caused by one pathogen, where disease severity is influenced by host and pathogen factors.

Two antigens that are common targets of the immune response to SARS-CoV-2 are the spike (S) glycoprotein, which is essential for both binding and entry into host cells, and the nucleocapsid (N) protein, involved in packaging the genomic material [4]. Antibodies to these antigens are induced after infection, and antibodies to S glycoprotein can be protective [5-7]. Indeed, the S glycoprotein is the sole SARS-CoV-2 viral antigen targeted by all current licensed vaccines [8]. After natural infection of non-vaccinated individuals, the appearance of antibodies to both of these antigens coincides with when severe disease develops. This means that substantial levels of viral antigen may still be present within the host for these antibodies to bind [9]. In contrast, in vaccinated, non-infected individuals, only anti-S antibodies are present at the time of pathogen encounter, and the levels of virus antigens at such times are likely to be lower than when antibodies become detectable during active infection.

After antigen binding by antibody, complement activation can occur through the classical pathway [10]. This cascade requires C1q binding to antibody and the generation of a C3 convertase derived in part from C4, through the production of C4b. This results in the cleavage of C3 and C5, with C3b and C5b forming a complex proximal to the site of antibody binding. The activation of the complement cascade may have positive or negative effects for the host associated with the timing of its activation and possibly the different pathways involved [11-13].

To improve our understanding of the relationship between SARS-CoV-2-specific antibodies and complement activation, we developed a solid phase C1q-binding assay and C4b, C3b and C5b complement deposition assays using S and N proteins from SARS-CoV-2. These studies identified differences in complement activation that were associated with the stage of infection in the host rather than the level of antibody detected.

## Methods

### Ethics and patient samples

Sera were obtained from distinct groups of subjects. Group 1: Non-vaccinated individuals without any reported COVID infection (NEG), with negative antibody responses to S and N using previously described assays [14, 15] Group 2: Individuals without evidence of infection (absence of anti-N antibodies), vaccinated 28-35 days previously with BNT162b2 vaccine (VACC). Group 3: Individuals without evidence of infection (absence of anti-N antibodies) who had received their second dose of BNT162b2 vaccine at least 28 days previously (DOUBLE VACC). Group 4: Non-vaccinated individuals recovered from a SARS-CoV-2 infection a minimum of 28 days previously (confirmed by antibody testing) who did not require hospitalisation (non-hospitalised convalescent patients, NHC). Group 5: Non-vaccinated convalescent, PCR confirmed SARS-CoV-2 infection patients who required ITU treatment, samples taken a minimum of 4 months after ITU discharge (ITU-CONV).

Ethical approval for obtaining samples for groups 1 -4 was provided by the London – Camden and Kings Cross Research Ethics Committee reference 20/HRA/1817. Ethical approval for obtaining samples for group 5 was provided by the North West ethics committee, Preston CIA UPH IRAS approval reference REC 20\NW\0240.

### Antigens used in this study

HEK293F cells were transiently transfected with SARS-CoV-2 HexaPro (GenBank: MN908947) to express metastable recombinant SARS-CoV-2 prefusion ectodomain. Engineered from the base construct 2P, HexaPro contains an extra 4 proline substitutions (residues 817, 892, 899, 942) in addition to those at residues 986 and 987 [16]. HexaPro exhibits antigenic properties equivalent to the 2P format [17]. HEK293F cells were cultured in Freestyle 293 Expression medium (Fisher Scientific) and maintained at a density of 0.2 × 10^6^ cells/mL at 37°C, 8% CO_2_ and 125 rpm shaking. Prior to transfection, two solutions of 25 mL Opti-MEM (Fisher Scientific) medium were prepared. Expression plasmid encoding SARS-CoV-2 HexaPro was added to the first solution to give a final concentration of 310 µg/L. To the other solution, 1 mg/mL pH7 polyethylenimine (PEI) max reagent was added to generate a ratio of 3:1 PEI max:plasmid DNA. Both solutions were combined and incubated at room temperature for 30 minutes. Cells were transfected at a density of 1 × 10^6^ cells/mL and incubated for 7 days at 37°C, 8% CO_2_ and 125 rpm shaking.

Cells were centrifuged at 3041g for 30 minutes at 4°C and supernatant was applied to a 500 mL Stericup-HV sterile vacuum filtration system (Merck) with a pore size of 0.22 µm. Purification of HexaPro S protein was undertaken using an ÄKTA Pure system (Cytiva). A 5 mL HisTrap Excel column (Cytiva) charged with Ni(II) was equilibrated using 10 column volumes (CV) of wash buffer (50 mM Na_2_PO_4_, 300 mM NaCl, pH 7). Supernatant was then loaded onto the column at a flow rate of 5 mL/min and washed with 10 CV of washing buffer containing 50 mM imidazole. Protein was eluted from the column in 3 CV of elution buffer (300 mM imidazole in washing buffer) and buffer exchanged to phosphate buffered saline (PBS) and concentrated using a Vivaspin column (MWCO 100 kDa) (Cytiva).

The nickel purified eluate was concentrated to 1 mL in PBS and injected into a Superdex 200 pg 16/600 column (Cytiva) to further purify trimeric S protein using size exclusion chromatography (SEC). The column was washed with PBS at 1 mL/min for 2 hours where fractions corresponding to the correct peak on the size exclusion chromatogram were collected and concentrated to ∼1 mL as above.

Nucleocapsid was generated as a recombinant protein from E. coli by the Protein Expression Facility at the University of Birmingham [15].

### Detection of antibodies specific to S and N

Antibody ELISAs were carried out as previously described [15], with 50 µl per dilution used. In brief, 96 well high-binding plates (Corning) were coated with 0.1 µg S or N protein in PBS and incubated overnight at 4°C. PBS-0.1% Tween 20 was used to wash plates 3 times, and between all subsequent steps. Plates were blocked with 2% (w/v) BSA in PBS-0.1% (v/v) Tween 20 for 1 hr at room temperature (RT). Serum was diluted 1:40 and incubated for 1 hr at RT. HRP-conjugated anti-human secondary antibodies were added for 1 hr at RT: anti-IgGAM, neat (EACONJ654, The Binding Site), anti-IgM clone AF6, 1:2000, anti-IgG1 clone MG6.41, 1:3000, anti-IgG3 clone MG5.161, 1:1000 (all produced at the University of Birmingham). Plates were developed for up to 20 minutes using 100 µl TMB Core (Bio-Rad) and the reaction was stopped with 50µl 0.2M H_2_SO_4_. Optical density (OD) was read at 450 nm using a SpectraMax ABS Plus plate reader.

### Solid phase C1q-binding assay

Plates were coated as above. Plates were washed three times with PBS-0.1% Tween 20 – this wash step was carried out between all subsequent steps. Blocking was carried out for 1 hr at RT with 2% BSA in PBS-0.1% Tween 20. Test serum was heat inactivated at 56□C for 30 minutes, before being diluted 1 in 5 with 2% BSA supplemented with 5 mM calcium chloride and 5 mM magnesium chloride. 50 µl was added to the antigen-coated plate and incubated for 1hr at 37□C. After washing, 50 µl COVID negative normal human serum (same source used throughout all assays, containing no detectable S or N specific antibodies as measured by IgGAM ELISA) at a dilution of 1:40 (in 2% BSA plus 5 mM calcium chloride and 5 mM magnesium chloride) was added to each well for 1hr at RT. 100µl of rabbit anti-C1q FITC antibody (Invitrogen PA5-16601) at a 1:200 dilution in PBS-0.1% Tween 20 was added and incubated at 37□C for 1 hr. HRP conjugated swine anti-rabbit (Dako P0399) at a 1:2000 dilution was then incubated for 1 hr. The assay was amplified using the Perkin Elmer ELAST amplification kit as per manufacturer’s instructions, with an optimised dilution of streptavidin, 1:800, incubated for 20 minutes. Plates were developed using 100 µl TMB Core (Bio-Rad) for 10 minutes, before being stopped with 50 µl 0.2MH_2_SO_4_. OD was measured as described above.

### C4b, C3b and C5b complement deposition assay

Microtiter plates were coated and washed as described above and blocked with Starting Block (ThermoFisher) for 10 min. Test serum was heat inactivated at 56□C for 30 minutes, before being diluted 1 in 5 with Starting Block supplemented with 5 mM calcium chloride and 5 mM magnesium chloride. 50 µl was added to the antigen-coated plate and incubated for 1hr at 37□C. After washing, 50 µl COVID negative normal human serum (same source used throughout all assays, containing no detectable S or N specific antibodies as measured by IgGAM ELISA) at a dilution of 1:40 (in 2% Starting Block plus 5 mM calcium chloride and 5 mM magnesium chloride) was added to each well for 1 hr at 37□C. The following anti-human monoclonal complement antibodies (100ul, diluted in PBS-0.1% Tween 20) were added and incubated at 37□C for 1 hr: mouse anti-C4b, 1:22,500 (Invitrogen, LF-MA0198); mouse anti-C3b, 1:10,000 (Invitrogen MA1-70053); mouse anti-C5b, 1:10,000 (Invitrogen DIA 011-01-02). HRP conjugated goat anti-mouse at a 1:4000 (Southern Biotech 1010-05) was then incubated at RT for 1 h. Plates were developed and read as described above.

### Statistics

Statistical analysis was carried out using GraphPad Prism 9.0. Kruskal-Wallis followed by Dunn’s post-hoc test for multiple groups was used to calculate p values. Statistical significance was accepted at P<0.05. Spearman correlation was carried out on the appropriate data sets.

## Results

### Anti-S, but not anti-N antibody responses differ between NHC and ITU-CONV patients

Total IgGAM antibody responses to trimeric S and N were assessed in five different groups: individuals without any reported COVID-19 infection (NEG); post first BNT162b2 vaccine, infection-naïve individuals (VACC); post second BNT162b2 vaccine, infection-naïve individuals (DOUBLE VACC); convalescing non-hospitalised patients (NHC) and convalescing patients who had been hospitalised and required ITU treatment (ITU-CONV). The VACC, DOUBLE VACC, NHC, ITU-CONV groups all had significantly higher anti-S glycoprotein IgGAM responses than the NEG group (Fig 1a), whereas IgGAM levels against N in the two convalescent groups were higher than the NEG, VACC and DOUBLE VACC groups (Fig 1b). Similar results were observed when specific IgG1 responses, an IgG isotype efficient at fixing complement, were assessed (Fig 1c). No differences in anti-S IgGAM and IgG1 antibody responses were observed between the VACC and patient groups. Anti-S IgGAM and IgG1 responses were higher in the ITU-CONV group compared to the NHC group, but no differences were observed for anti-N responses in these two groups (Fig 1d). Modest IgM and IgG3 responses to S and N were detected in only a few individuals (Supp Figs *1a and b*).

**Fig 1:**
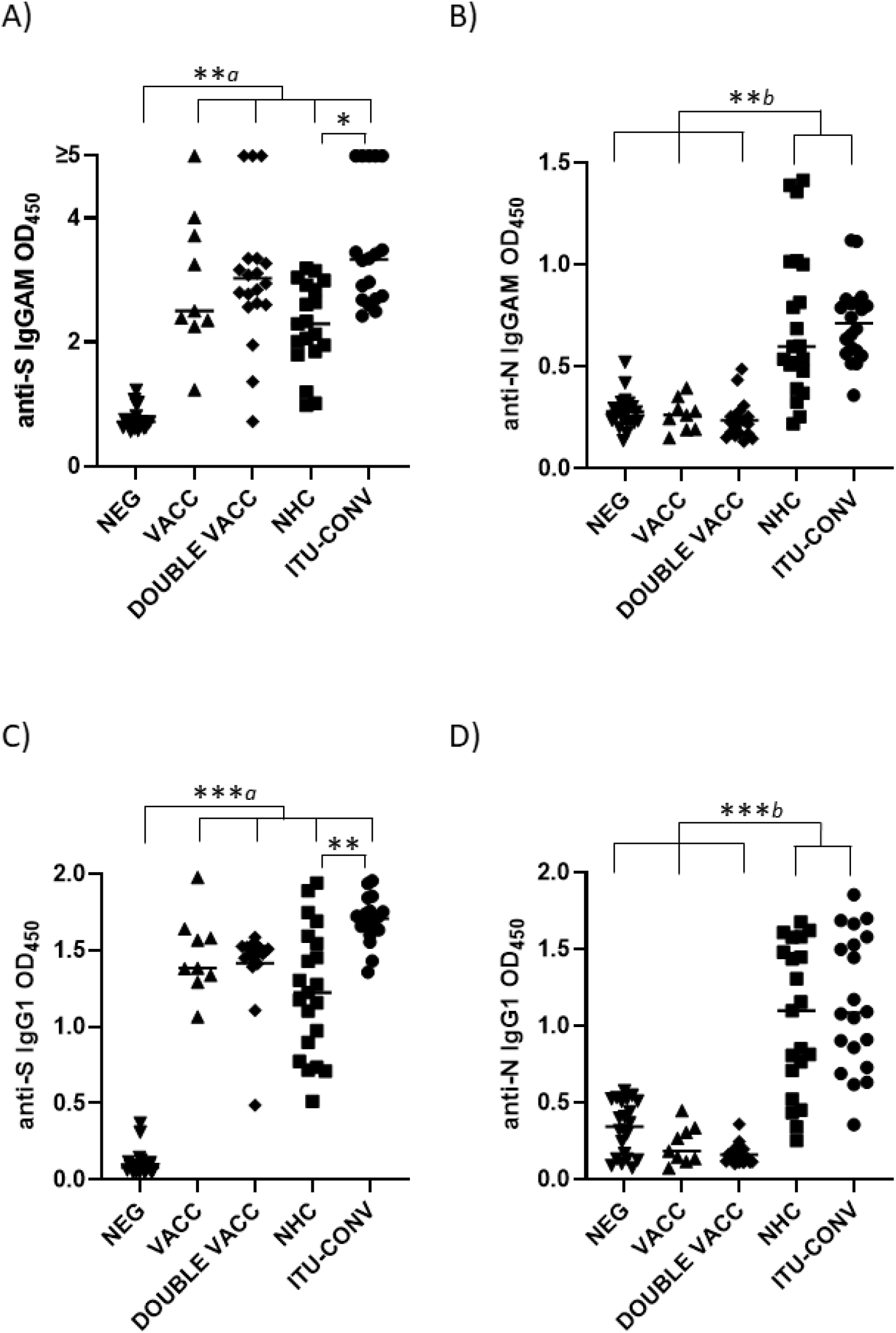
Anti-S, but not anti-N antibody responses differ between NHC and ITU-CONV patients. Using an ELISA against 0.1ug S (a, c) or N (b,d) with HRP-conjugated IgGAM or IgG1 secondary antibodies, GAM and IgG1 levels were assessed in the following subject groups: COVID-19 negatives (NEG, n ≥20), COVID-19 naïve one month post first BNT162b2 vaccine (VACC, n = 9), COVID-19 naïve one month post second BNT162b2 vaccine (DOUBLE VACC, n = 19), COVID-19 positive non-hospitalised convalescents (NHC, n ≥ 19) and COVID-19 positive convalescents who required ITU treatment (ITU-CONV, n ≥ 18) Kruskal-Wallis with Dunn’s multiple comparisons test was used to test significance. *a* indicates that the four groups bracketed (VACC, DOUBLE VACC, NHC and ITU-CONV) were individually significantly different to the NEG group; *b* indicates that NHC and ITU-CONV are independently significantly different to NEG, VACC and DOUBLE VACC. *** p < 0.001, ** p < 0.01, * p < 0.05. Bars represent median values for each group.

### C1q binding *in vitro* correlates with levels of S- and N-specific IgGAM and IgG1 antibodies

To determine if the complement protein C1q can bind to SARS-CoV-2-specific immunoglobulins *in vitro* we developed a solid phase C1q-binding assay. In these antigen-specific assays, the test serum from COVID-19 patients or vaccinees is heat-inactivated and standardisation of complement is provided by using sera from non-infected, non-vaccinated subjects. Results from this assay showed that C1q binding mirrored IgG1 levels for both S and N antigens, with the lowest signals for S seen in the NEG group (Fig 2a), and for N in the NEG, VACC and DOUBLE VACC groups (Fig 2b). No difference in C1q binding was observed between sera from the two convalescent groups (Figs 2 a and b). Plotting IgG1 responses against C1q responses shows a positive correlation between the amount of IgG1 antibody and the amount of C1q binding detected (Fig 2c and d). Therefore, C1q binding reflects the serological response to both antigens.

**Fig 2:**
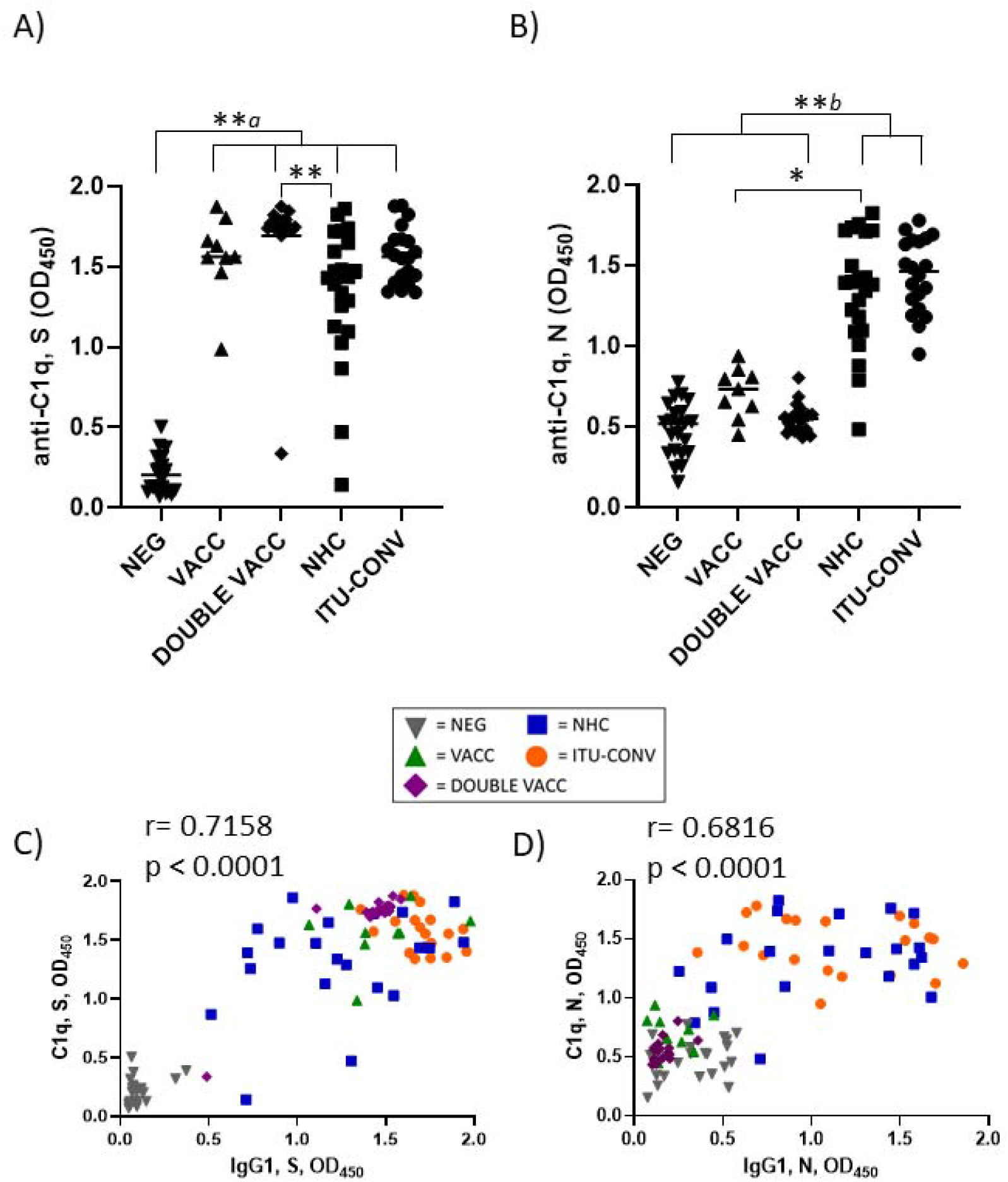
C1q binding to S and N correlates with IgG1 responses. Using an ELISA against 0.1ug S (a) or N (b) with an anti-C1q secondary antibody, followed by an HRP-conjugated tertiary, and the ELAST amplification kit, C1q binding was measured. Correlations of IgG1 OD and C1q OD against S (c) and N (d). NEG, n= 22. VACC, n =9. DOUBLE VACC n = 19. NHC, n ≥ 21. ITU-CONV n = 20. Kruskal-Wallis with Dunn’s multiple comparisons test was used. *a* indicates that the four groups bracketed were individually significantly different to the NEG group; *b* indicates that NHC and ITU-CONV are independently significantly different to NEG and DOUBLE VACC. ** p < 0.01, * p <0.05. Bars represent median values for each group. Spearman correlation was used to assign r and p values.

### Deposition of C4b, C3b and C5b varies dependent upon antigen tested and subject group

To determine whether C1q binding reflected downstream activation of the complement cascade, we examined whether complement breakdown products could be detected. Deposition of C4b, a major component of the classical pathway C3 convertase, and the effector molecules C3b and C5b were assessed. In the absence of S or N-specific antibodies, C4b, C3b and C5b breakdown products were not detected, but they were detected in the presence of specific antibodies, indicating involvement of the classical complement pathway (Fig 3). When S was used as the assay antigen, the highest median levels of C4b, C3b and C5b deposition detected were in the DOUBLE VACC and ITU-CONV groups (Fig 3a), whereas the VACC and NHC groups showed similar lower levels of downstream activation. The median levels of C4b, C3b and C5b deposition detected when N was used as the assay antigen were similar between the NHC and ITU-CONV groups (Fig 3b). Therefore, in this assay differences in complement activation can be detected dependent upon what patient group and antigen were examined.

**Fig 3:**
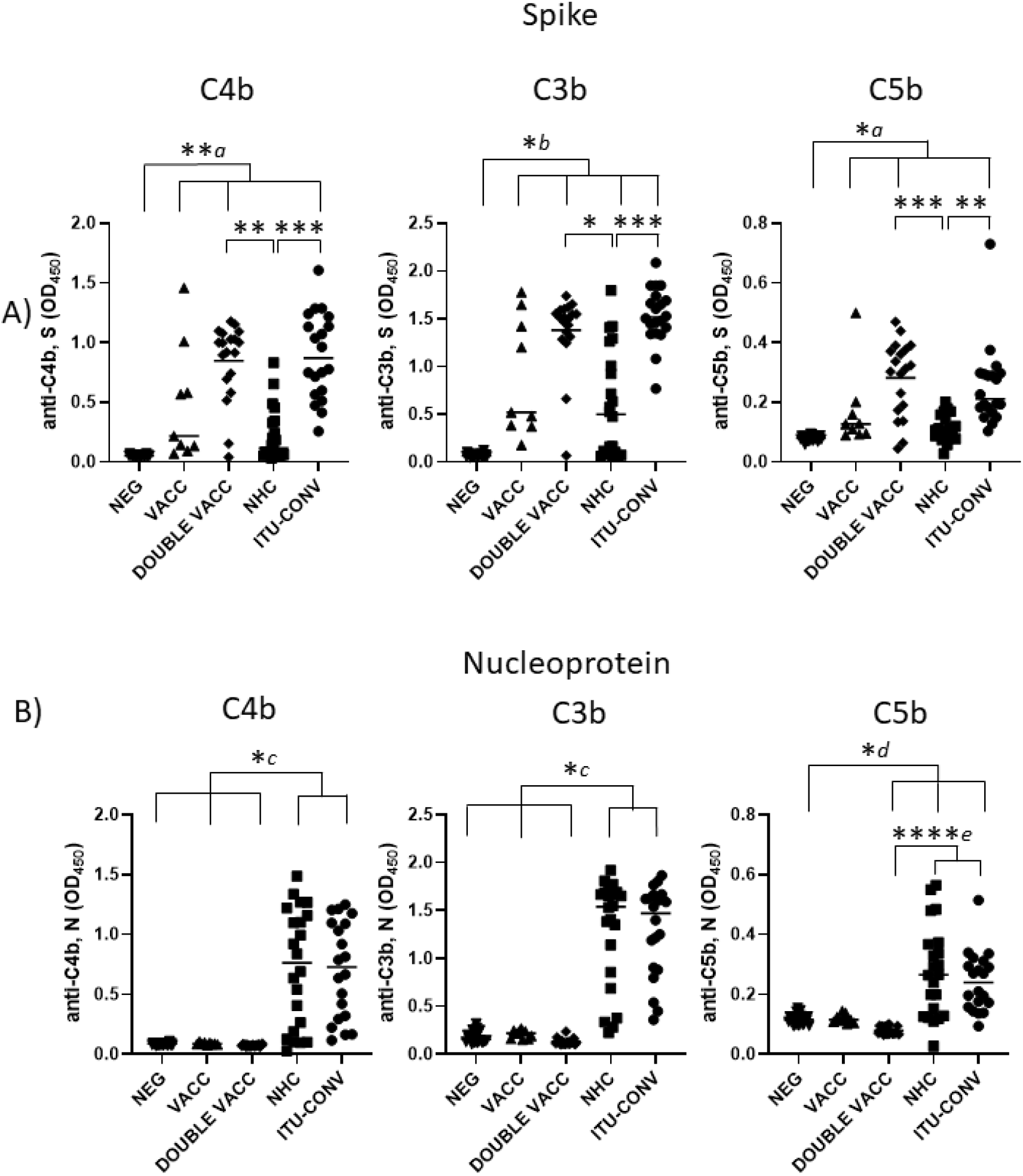
C4b, C3b and C5b show antigen and subject status-dependent variability. Using an ELISA against 0.1ug S (a) or N (b) with either anti-C4b, C3b or C5b secondary antibody, followed by an HRP-conjugated tertiary, downstream complement binding was measured. NEG, n= 22. VACC, n =9. DOUBLE VACC, n = 19. NHC, n = 22. ITU-CONV n = 20. Kruskal-Wallis with Dunn’s multiple comparisons test was used. *a* and *b* indicate that the groups bracketed were individually significantly different to the NEG group; *c* indicates that NHC and ITU-CONV are independently significantly different to NEG, VACC and DOUBLE VACC; *d* indicates that DOUBLE VACC, NHC and ITU-CONV are all significantly different to NEG; *e* indicates that NHC and ITU-CONV are both significantly different to DOUBLE VACC. **** p < 0.0001, *** p < 0.001, ** p < 0.01, * p <0.05. Bars represent median values for each group.

### Downstream complement activation associates with threshold anti-S IgG1 responses

In contrast to the linear association between anti-S IgG1 and C1q detection, there was a non-linear association between IgG1 and C4b, where C4b was only detectable beyond a threshold level of IgG1. Although there was a correlation between C4b and IgG1 to N, such a threshold response was not observed (Fig 4a). This threshold response for IgG1 to S but not N was also observed if C3b or C5b were plotted against IgG1 (Supp Figs *2a* and *2b*). When correlations were performed for C1q vs C4b (Fig 4b), C4b vs C3b (Fig 4c) and C3b vs C5b (Fig 4d) for both antigens, then a more linear relationship was observed. This suggests that in this assay higher levels of anti-S IgG1 are needed to activate downstream complement components than for N.

**Fig 4:**
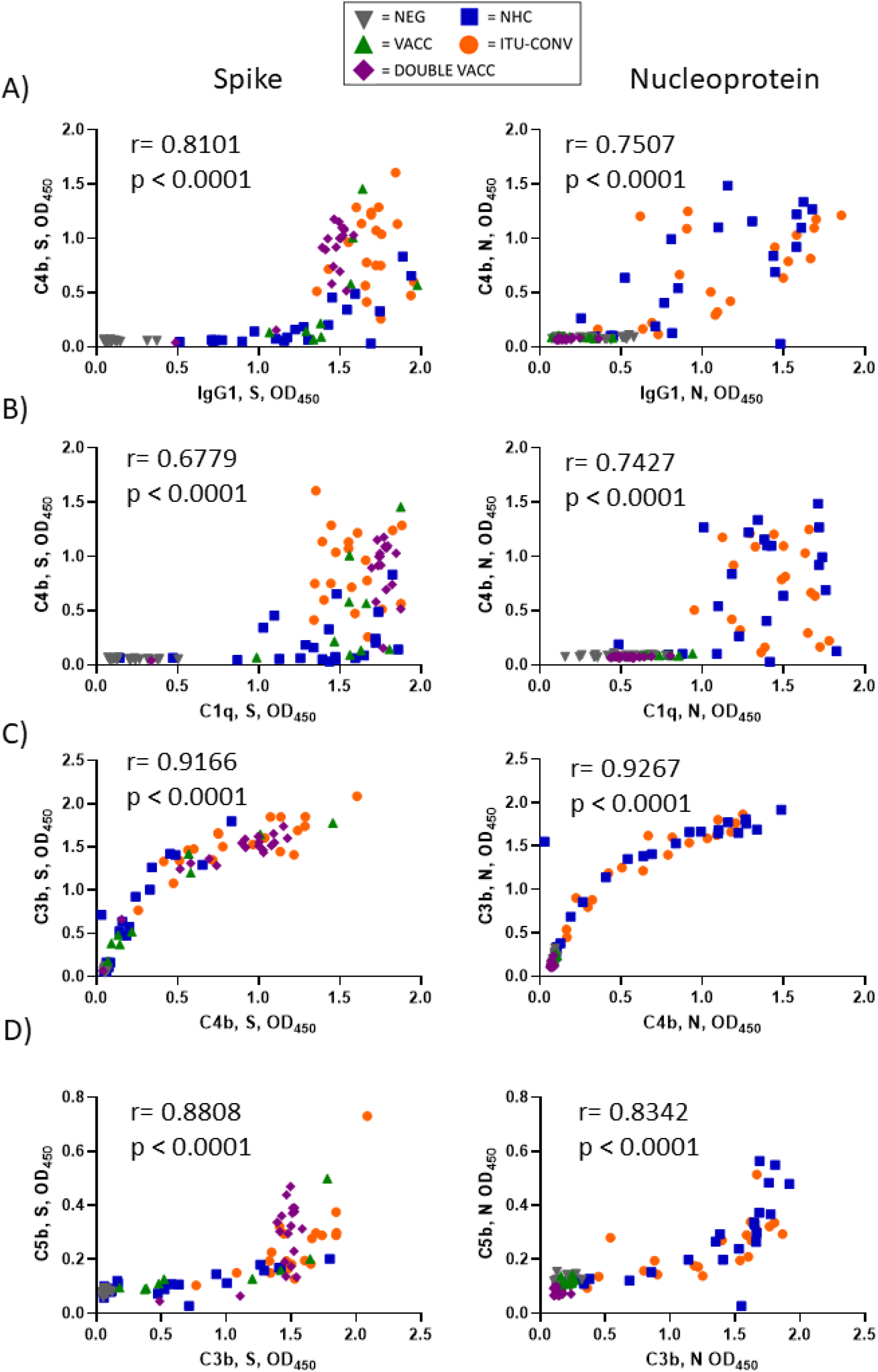
C4b responses demonstrate a threshold response to IgG1 levels against S but not N. All further downstream complement components correlate linearly. Correlations of data obtained in Fig 1c and 1d, Fig 2a and 2b and Figs 3a and 3b were plotted. XY pairs, n ≥ 91. Spearman correlation was used to assign r and p values.

## Discussion

Here we show (i) that antibodies to S and N can activate the classical complement cascade and (ii) the level of activation of the cascade detected can vary dependent upon the antigen and source of the antibodies. Therefore antibodies to two different antigens within the same pathogen can activate complement, albeit at levels that depend on the severity of COVID-19. Since antibody binding to both antigens resulted in the binding of C1q, the ability to initiate the cascade is not limiting to these antigens in this assay. For sera from the two convalescent groups, greatest variability was detected downstream of C1q binding and was somewhat dependent upon the nature of the antigen. The reasons why this could occur are unclear, but reasons may include intrinsic differences in the antigens themselves. For instance, S is a trimeric protein and the approximately 420 kDa trimer is substantially larger than N, which is approximately 46 kDa. This difference may influence how IgG binds to the antigen and affects IgG hexamerisation [18]. More investigation is needed to understand the relationship between antigen, antibody and complement activation more fully.

The results from these studies lead to further hypotheses to test. For instance, it could be hypothesised that ITU subjects have greater activation of the classical complement cascade during infection and this compounds their disease. Alternatively, since all these subjects survived severe COVID-19 infections it could be hypothesised that the activation of complement is associated with a beneficial outcome. As we did not have sera from individuals who died this is not testable here. One caveat in this argument is that exogenous sources of complement in the form of sera from non-infected individuals were used in these studies and that patients own sera may differ in potency. This was not assessed here as the focus was on antibody-mediated activation of complement. Additionally, these assessments were made using sera from patients who were infected or immunized weeks previously and the antibodies present may not reflect the antibodies present at the time of infection.

Certainly, it could be expected that the affinity of the antibodies would increase over time. One striking feature was the variability in the anti-C4b/C3b response detected in the VACC group. It is unclear why this is the case, but it could simply be that there is variability within the wider population in the ability to activate complement downstream.

The complement cascade has been reported to be activated through multiple pathways after SARS-CoV-2 infection [19-21]. Amongst these, the engagement of the classical pathway is distinct to the non-antibody-dependent pathways due to the potential multiple roles antibody can play during the course of infection. If induced whilst an infection is ongoing, then the activation of the complement cascade by antibody could worsen disease, particularly as antibody responses become detectable concomitant with risk of severe disease. This could happen either through enhanced inflammation, such as observed during acute respiratory distress syndrome, or through enhancing the complications of thrombosis and coagulopathy after infection [22, 23]. Moreover, antibodies are induced to multiple SARS-CoV-2 antigens and as we show, antibodies to S and N proteins have the capacity to activate complement in vitro. Balancing this, positive roles for antibody-mediated complement activation have also been proposed during active infection and vaccination [12]. Potentially, the most valuable contribution antibody-mediated complement could make to protection is in vaccinated individuals. Here, antibody-mediated activation of complement may be more beneficial for the host because it is contributing to control of infection when the pathogen burden is relatively low and less likely to provoke severe inflammatory responses.

In summary, we have identified activation of the classical complement pathway after vaccination against SARS-CoV-2, or after infection with this pathogen. The variability in the responses we detect will help us understand how complement is activated in the presence of antibodies and how this may contribute to protection and harm in those who encounter this pathogen.

## Data Availability

All data produced in the present work are contained in the manuscript

## Acknowledgments

We would like to thank the staff of the Clinical Immunology Service, University of Birmingham for their invaluable work in sample collection and processing. We also gratefully acknowledge the University of Birmingham Protein Expression Facility for the production of nucleoprotein. We thank Jason McLellan for providing the expression plasmid for HexaPro.

## Figure legends

**Supplementary figure 1:**
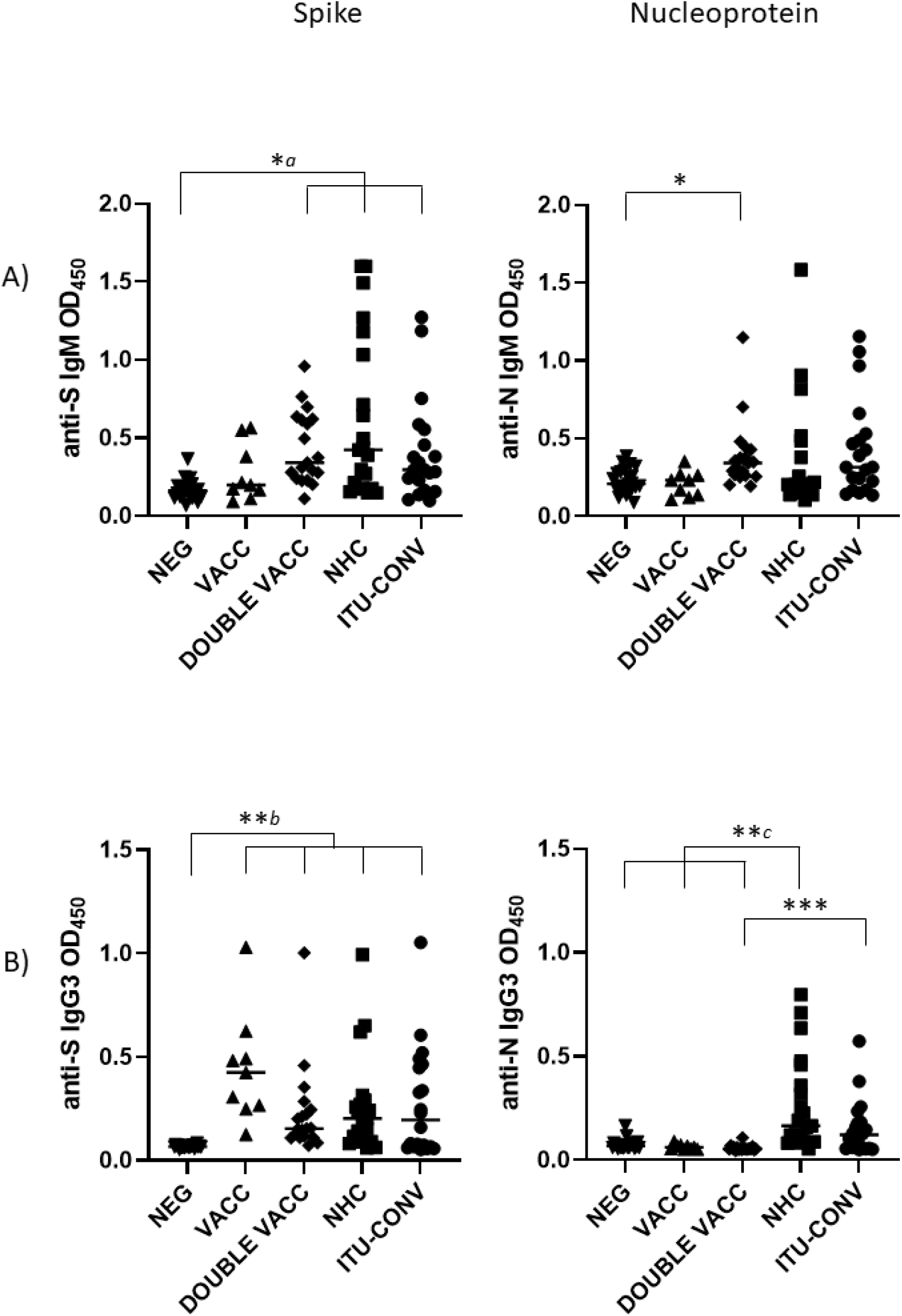
Convalescent and double vaccinated individuals have higher levels of IgM to S than non-infected individuals, but responses to N do not differ significantly. Levels of IgG3 are higher in convalescent and vaccinated groups than negatives to S, but only convalescent groups have higher levels to N. Serological responses were assessed by ELISA using HRP labelled a) anti-IgM and b) IgG3 against 0.1ug purified S or N. Kruskal-Wallis with Dunn’s multiple comparisons test was used to test significance. *a* and *b* indicate that the groups bracketed were individually significantly different to the NEG group; *c* indicates that NEG, VACC and DOUBLE VACC are independently significantly different to NHC. *** p < 0.001, ** p < 0.01, p < 0.05. Bars represent median values for each group. NEG, n = 22. VACC, n = 9. DOUBLE VACC, n = 19. NHC, n = 21. ITU-CONV, n = 20.

**Supplementary figure 2:**
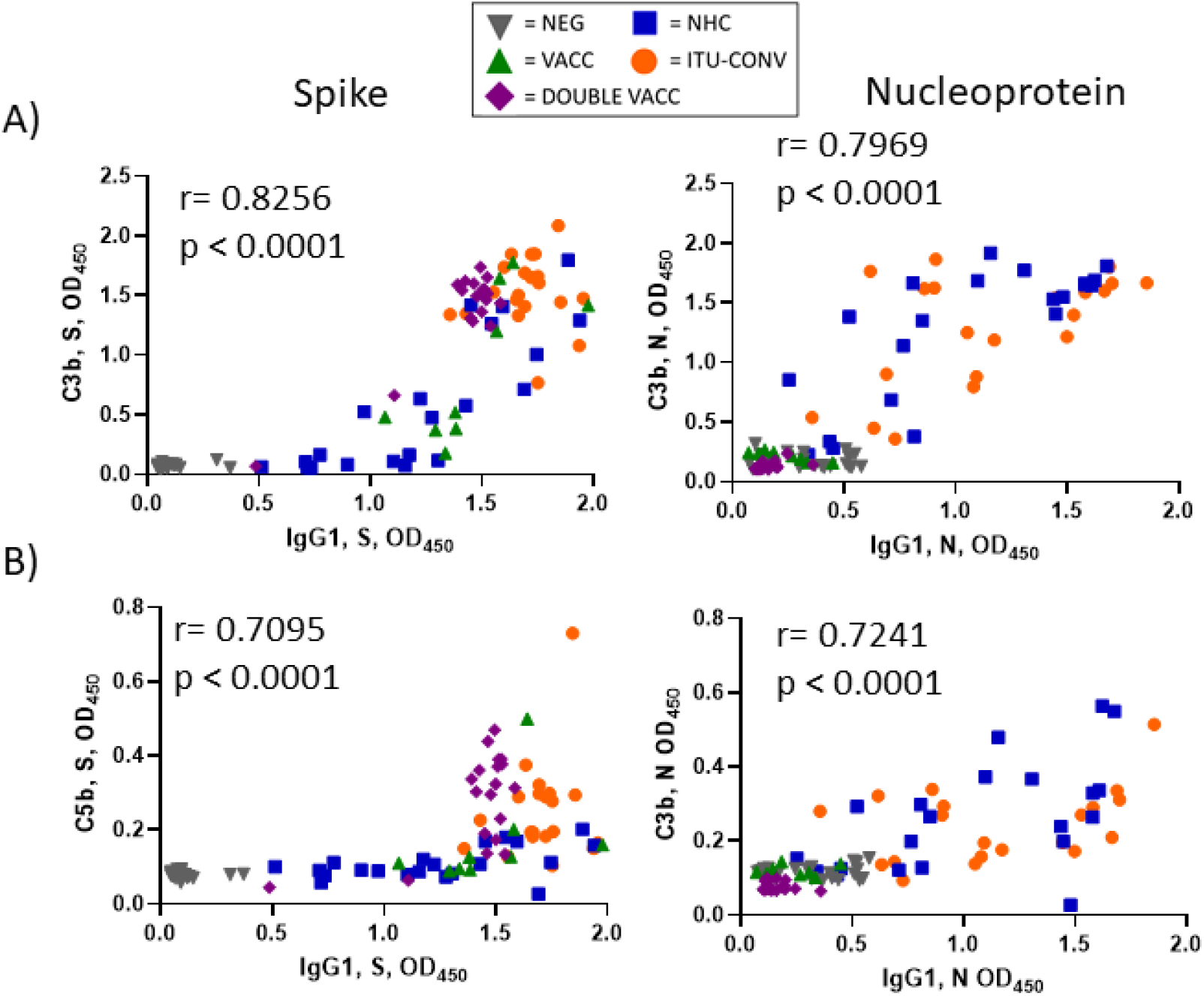
C3b and C5b responses demonstrate a threshold response to IgG1 levels against S but not N. Correlations of data obtained in Figs 1c and 1d, and Figs 3a and 3b were plotted. XY pairs, n = 91. Spearman correlation was used to assign r and p values.

